# Can Demographic and Health Surveys (2007–2024) Capture Alcohol Use Trends in Zambia?

**DOI:** 10.64898/2026.08.18.26360685

**Authors:** Shadreck Habbanti, Picket Munkombwe, Cosmas Zyambo

## Abstract

**Background:** Alcohol is a leading modifiable risk factor for non-communicable disease. Zambia’s National Alcohol Policy and the World Health Organization’s target of a 10% relative reduction in the harmful use of alcohol both require that the trend be monitored. Alcohol items appear in four rounds of the Zambia Demographic and Health Survey (ZDHS), and those rounds are widely treated as a trend series, although whether they are one has never been tested.

**Methods:** Secondary analysis of ZDHS 2007, 2013–14, 2018 and 2024 (women aged 15–49, men aged 15–59). A direct current-use item is available in three rounds, with three different instruments for men and two for women. Four identification strategies were applied in ascending order of assumption: nesting bounds, which exploit the fact that a seven-day window falls within a thirty-day window and that within undated current status; restriction to the fieldwork months common to both rounds; a lifetime-use analogue available in 2024; and an instrument-constant partner-report series available in all four rounds, validated by linking each woman to her co-resident husband. Estimation throughout was design-based.

**Results:** The conventional series suggests a fall in current drinking among men from 42.0% (95% CI 39.9–44.1) in 2007 to 28.2% (95% CI 27.0–29.3) in 2024, and among women from 11.1% (95% CI 9.9–12.4) to 8.8% (95% CI 8.0–9.6). Neither change is sign-identified. Placed on a common thirty-day basis with matched fieldwork months, the 2013–14 to 2024 change lies between -7.6 and +1.2 percentage points for men and between -0.2 and +3.7 for women. The instrument-constant proxy fell from 53.7% in 2007 to 37.7% in 2018, then plateaued at 37.0% in 2024; a constant-decline model is rejected (Q = 15.9, 2 df, p = 0.0003). Sensitivity of the proxy against husbands’ own reports fell from 86.0% to 66.8%. The 2024 cross-section is unaffected and is reported in full.

**Conclusions:** These data do not establish the apparent national decline in alcohol use. Differences in reported prevalence across ZDHS rounds substantially reflect instrument change, reference-period shift, fieldwork seasonality and decay in proxy reporting. On present evidence Zambia cannot monitor its alcohol commitments from national survey data. Trend monitoring would require a consistent alcohol module restored to the questionnaire, an occasion-based heavy-drinking item, and the reporting of fieldwork month.

## Introduction

Alcohol accounts for a substantial share of the global burden of non-communicable disease and injury, tracked through adult per-capita consumption and heavy episodic drinking (1,2); harm is graded by the amount consumed, not by whether a person drinks at all (3). Regulation of price, availability and marketing is among the World Health Organization’s "best buys" (4). Zambia has legislated accordingly, through the Liquor Licensing Act of 2011 and the National Alcohol Policy (5,6), and has committed to a 10% relative reduction in harmful alcohol use (7); sub-Saharan Africa is not on track to meet that target (8). Whether the trend can be observed at all is rarely asked.

Zambia’s observational base is thin. The most recent regional synthesis finds comprehensive data scarce and cannot place the underlying studies on a common reference frame (9); the 2017 STEPS survey gives an instrument-consistent binge measure but was never repeated (10); the HIV impact assessments field a harmonized screen but no series (11). Four ZDHS rounds carry alcohol items, the only repeated national source, and in practice are treated as a trend series. Two features of the Zambian market make that risky: traditional, home-brewed and illicitly distilled beverages dominate consumption (12,13), so an item naming one beverage category measures something narrower than alcohol, and prevalence depends heavily on question form, with twelve operational definitions of current drinking in use (14) and Zambian estimates shifting by up to 60% with item wording (15).

Secondary analyses of DHS alcohol items commonly acknowledge round differences in a limitations paragraph and estimate a trend regardless. This paper asks first whether a comparable series exists, and where it does not, what the rounds can still support. The answer is largely negative but actionable: it names the instrument features that would have to be improved. One positive finding survives. The partner-report items, worded identically in all four rounds (16,17), supply a series spanning even the round with no direct item; it falls steeply, then flattens.

## Methods and Measurements

This is a desk-based secondary analysis of de-identified individual recodes from four ZDHS rounds: 2007, 2013–14, 2018 and 2024 (18,19). Women aged 15–49 and men aged 15–59 were interviewed in every round. A direct own-use item exists for 71,330 respondents across three rounds (33,849 men and 37,481 women); 2018 contributes no direct outcome and enters only through the proxy analysis.

### The instrument inventory

The alcohol items differ across rounds in three ways: beverage frame, the 2007 men’s item asking about beer alone; reference period, 2007 and 2013–14 asking an undated status question with days drunk in the last week where 2024 asked days drinking in the past thirty; and item presence, 2018 carrying no direct own-use item for either sex. Table 1 sets out the inventory.

**Table 1.** Alcohol items by ZDHS round and sex.

| Round | Women's item | Men's item | Reference period | Note |
| --- | --- | --- | --- | --- |
| 2007 | s1012a / s1012b | sm812 / sm813 | Undated status; days in last week | Men's item is beer only |
| 2013–14 | s1007a / s1007b | sm811a / sm811b | Undated status; days in last week | All alcohol, both sexes |
| 2018 | none | none | — | No direct own-use item |
| 2024 | v485a / v485b | mv485a / mv485b | Days in past 30; drinks per day | Codes 0, 95, 96 distinguish none this month, near-daily, never |
| All four | d113, d114 | — | Undated status, partner-reported | Instrument-constant; universe widened in 2024 |

### Survey design and estimation

All estimates are survey-weighted, with a stratified, cluster-robust linearized variance estimator on primary sampling units; partner-item estimates use the domestic-violence module weight, correcting for one-woman-per-household selection and module non-response. Confidence intervals are normal-approximation intervals on the percentage scale. Analyses ran in Python 3.12; the full pipeline is deposited and citable.

### Identification strategies

Where a parameter cannot be recovered without assumptions the data cannot support, bounding it is the appropriate response (20,21). In DHS data from Zambia and neighbouring countries, imputation under missing-at-random produced seriously biased prevalence once that assumption was mildly violated; graded assumptions returned usable bounds (22). The four strategies below follow that logic, each asking more of the data than the last (23).

Strategy A exploits nesting: a seven-day window sits inside a thirty-day window, itself inside undated current status, so the two earlier rounds bracket the figure 2024 reports directly; estimates separate cleanly by reference period in sub-Saharan African data (24,25), and a shortened frame depresses DHS estimates through omission and displacement of events (26). Strategy B restricts both rounds to their common fieldwork months, a recent-window measure being seasonal and calendar month a first-order source of variation in DHS estimates (27). Strategy C separates, within the 2024 item, never-drinkers from those who did not drink that month, a lifetime measure and upper bound on current status. Strategy D links each module-eligible woman to her co-resident husband to test the partner items directly (28,29); within-couple concordance is high in southern Africa (30), with sensitivity and specificity estimated against the husband’s own report (31,32). The partner analysis covers ever-married women, the universe every round asked. No modelled seven-day equivalent and no projection are reported, the input trend having proved not to be sign-identified.

### The 2024 cross-section

A single round carries no comparability problem, so the 2024 data are analysed directly: prevalence across sociodemographic groups, the never, former and current decomposition, drinking days in the past month and usual drinks per drinking day. The share reporting five or more drinks (men) or four or more (women) on a usual day is reported as usual heavy quantity, not heavy episodic drinking, since the item asks about a typical occasion. Determinants come from a 2024-only logistic model fitted separately by sex; a pooled model is retained only as a sensitivity analysis.

## Results

### Participants

Interviews in the 2024 round covered 12,585 men aged 15–59 and 13,951 women aged 15–49, each giving a direct alcohol response. Table 2 sets out their characteristics on every variable used anywhere in this paper and supplies the fitted terms for the regression in Tables 9 and 10, with categories and reference levels identical in both. Missingness never exceeds 0.3% of records on any variable in any round, so the analysis runs complete-case.

**Table 2.**
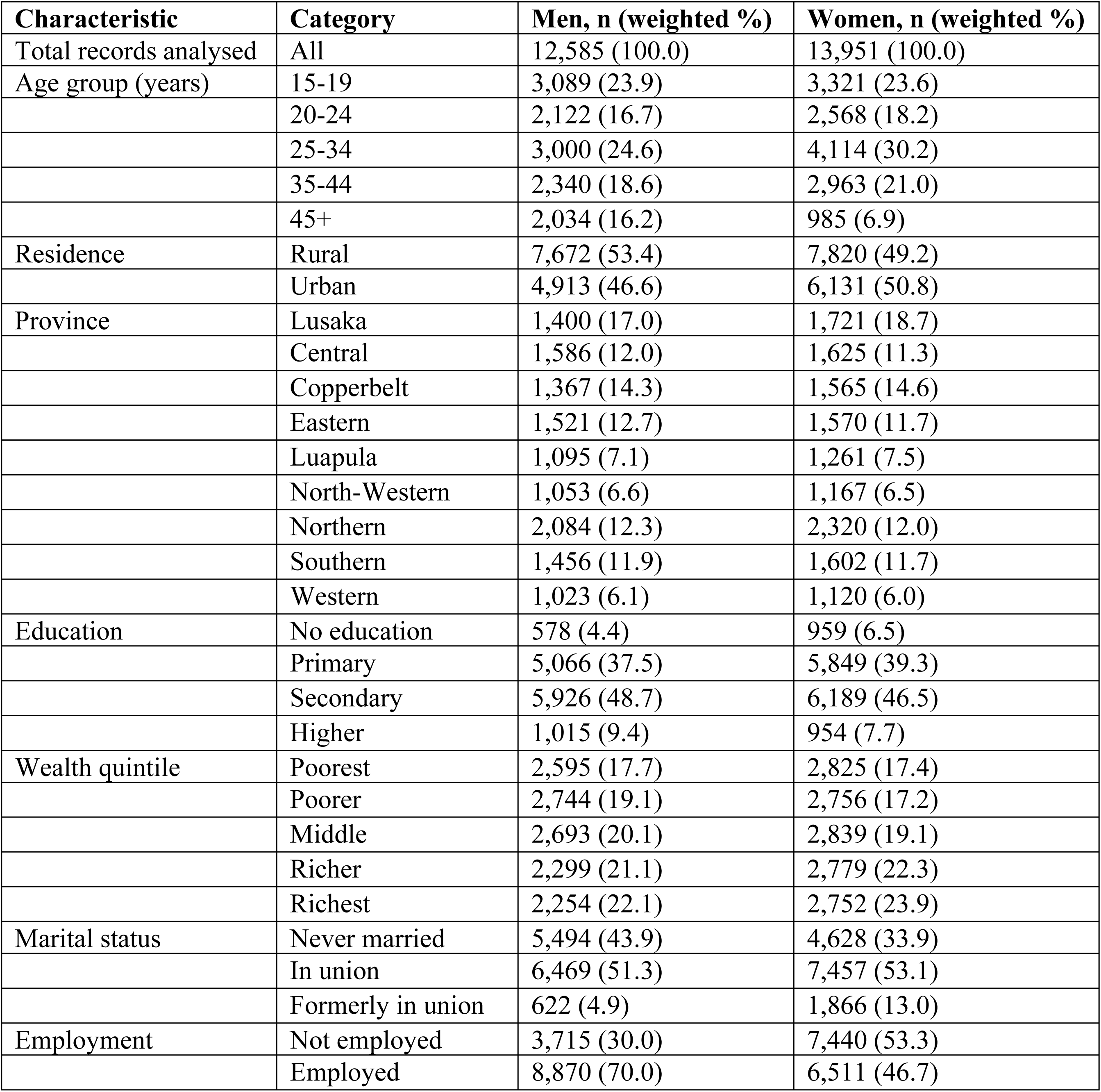
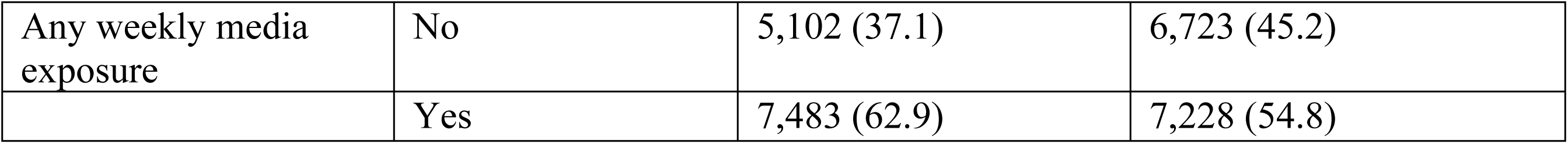
Characteristics of all records analysed, ZDHS 2024, by sex. Percentages are survey-weighted, counts unweighted; regression reference categories are the first row of each characteristic.

**Table 3.** Composition of the analytic sample by round and sex. The 2018 round contributes records but no direct alcohol item.

| Round | Sex | Records | With a direct item | % urban | % in union |
| --- | --- | --- | --- | --- | --- |
| 2007 | Men | 6,500 | 6,494 | 43.2 | 55.8 |
|  | Women | 7,146 | 7,127 | 42.1 | 61.6 |
| 2013 | Men | 14,773 | 14,770 | 46.1 | 55.1 |
|  | Women | 16,411 | 16,404 | 46.2 | 60.1 |
| 2018 | Men | 12,132 | 0 | 44.1 | 53.0 |
|  | Women | 13,683 | 0 | 46.6 | 55.9 |
| 2024 | Men | 12,585 | 12,585 | 46.6 | 51.3 |
|  | Women | 13,951 | 13,950 | 50.8 | 53.1 |

**Table 4.**
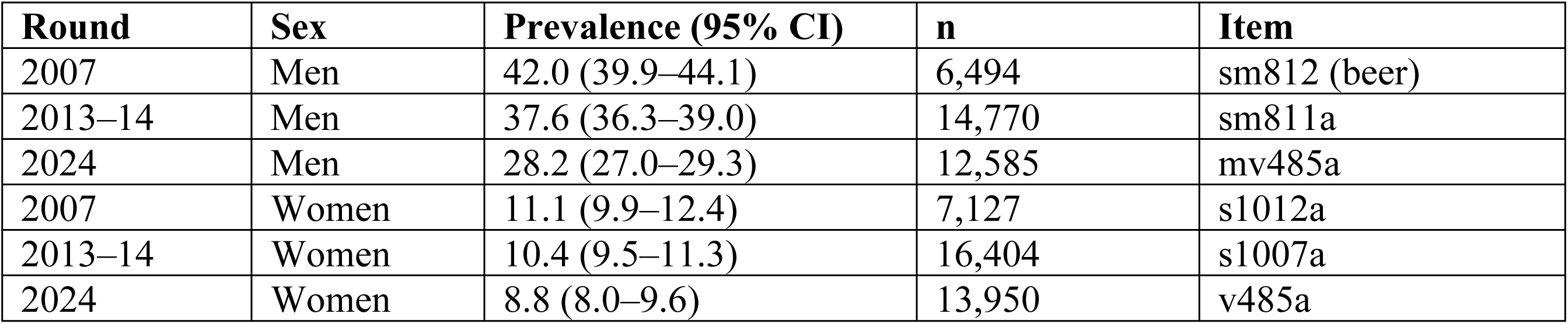
Design-based prevalence of current alcohol use, by round and sex, with the item used for each estimate.

| Round | Sex | Prevalence (95% CI) | n | Item |
| --- | --- | --- | --- | --- |
| 2007 | Men | 42.0 (39.9–44.1) | 6,494 | sm812 (beer) |
| 2013–14 | Men | 37.6 (36.3–39.0) | 14,770 | sm811a |
| 2024 | Men | 28.2 (27.0–29.3) | 12,585 | mv485a |
| 2007 | Women | 11.1 (9.9–12.4) | 7,127 | s1012a |
| 2013–14 | Women | 10.4 (9.5–11.3) | 16,404 | s1007a |
| 2024 | Women | 8.8 (8.0–9.6) | 13,950 | v485a |

**Table 5.** Identification intervals for the 2013–14 to 2024 change on a common thirty-day basis, before and after restriction to common months.

| Sex | Fieldwork scope | 2013–14 bracket | 2024 observed | Implied change |
| --- | --- | --- | --- | --- |
| Men | All fieldwork | 30.5–37.6% | 28.2% | -9.5 to -2.3 pp |
| Men | Jan-Apr only | 26.9–35.8% | 28.1% | -7.6 to +1.2 pp |
| Women | All fieldwork | 6.1–10.4% | 8.8% | -1.6 to +2.7 pp |
| Women | Jan-Apr only | 4.8–8.7% | 8.6% | -0.2 to +3.7 pp |

**Table 6.** Partner-reported alcohol indicators, identical wording in all four rounds, ever-married women.

| Round | Partner drinks alcohol | Partner often drunk | Ever-married women (n) |
| --- | --- | --- | --- |
| 2007 | 53.7% (51.7–55.6) | 19.9% (18.3–21.5) | 4,235 |
| 2013–14 | 46.5% (44.9–48.1) | 15.3% (14.2–16.4) | 9,410 |
| 2018 | 37.7% (35.5–39.9) | 12.7% (11.6–13.8) | 7,358 |
| 2024 | 37.0% (35.4–38.6) | 12.9% (11.8–13.9) | 7,272 |

**Table 7.**
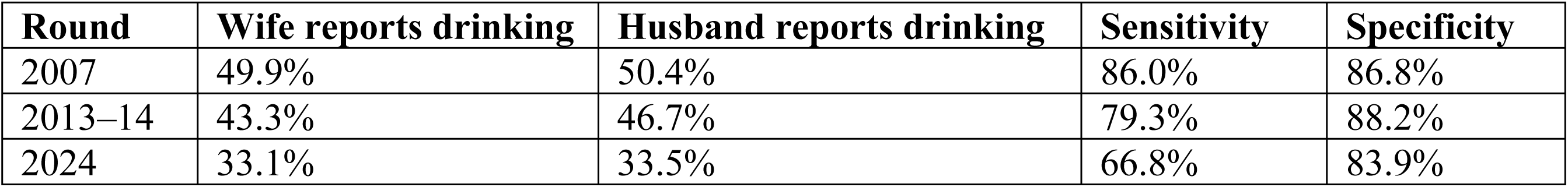
Agreement between women’s reports of partner drinking and husbands’ own reports, co-resident couples.

**Table 8.** Summary of what the four ZDHS rounds do and do not establish.

| Claim | Basis | Verdict |
| --- | --- | --- |
| Men declined, 2007 to 2013–14 | Matched 7-day items, but beer versus alcohol | Not interpretable |
| Women declined, 2007 to 2013–14 | Matched items and matched window | Identified |
| Men declined, 2013–14 to 2024 | Nesting bounds, season-matched | Not identified |
| Women declined, 2013–14 to 2024 | Nesting bounds, any scope | Not identified |
| Partner drinking declined to 2018 | Instrument-constant proxy | Identified, part measurement decay |
| Decline continued after 2018 | Instrument-constant proxy | Rejected |

**Table 9.**
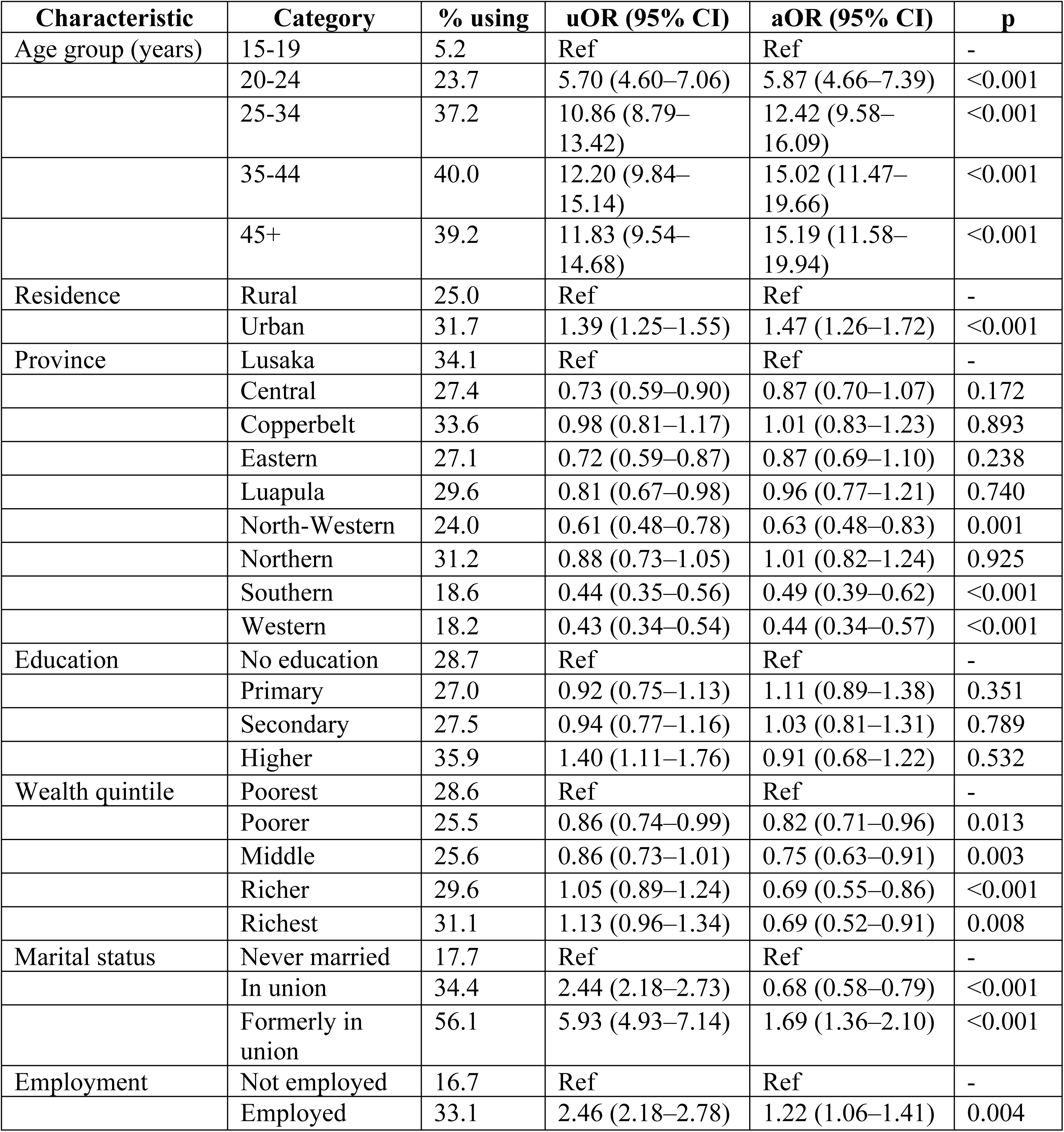
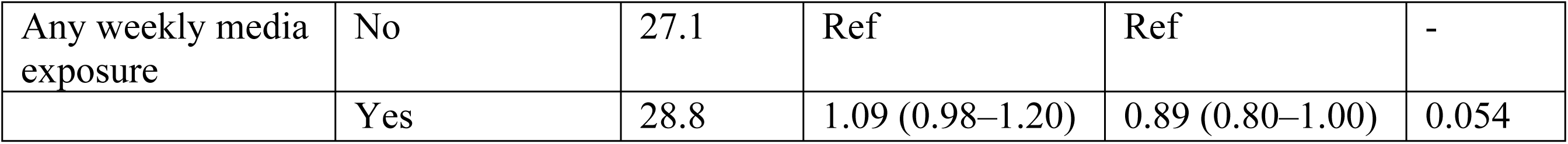
Determinants of current alcohol use among men, ZDHS 2024 (n = 12,585). Unadjusted and adjusted odds ratios from design-based logistic regression; the adjusted model contains every characteristic shown. Reference categories carry an odds ratio of 1.00; p-values are for the adjusted model.

**Table 10.** Determinants of current alcohol use among women, ZDHS 2024 (n = 13,950). Unadjusted and adjusted odds ratios from design-based logistic regression; the adjusted model contains every characteristic shown. Reference categories carry an odds ratio of 1.00; p-values are for the adjusted model.

| Characteristic | Category | % using | uOR (95% CI) | aOR (95% CI) | p |
| --- | --- | --- | --- | --- | --- |
| Age group (years) | 15-19 | 2.8 | Ref | Ref | - |
|  | 20-24 | 7.0 | 2.67 (1.97–3.62) | 2.73 (2.00–3.75) | <0.001 |
|  | 25-34 | 10.7 | 4.23 (3.18–5.62) | 4.41 (3.22–6.03) | <0.001 |
|  | 35-44 | 13.3 | 5.40 (4.02–7.25) | 5.82 (4.13–8.19) | <0.001 |
|  | 45+ | 12.1 | 4.86 (3.41–6.92) | 5.07 (3.40–7.57) | <0.001 |
| Residence | Rural | 5.0 | Ref | Ref | - |
|  | Urban | 12.5 | 2.71 (2.23–3.28) | 1.62 (1.25–2.09) | <0.001 |
| Province | Lusaka | 12.7 | Ref | Ref | - |
|  | Central | 7.7 | 0.57 (0.42–0.79) | 0.94 (0.69–1.27) | 0.671 |
|  | Copperbelt | 15.3 | 1.25 (0.95–1.63) | 1.38 (1.07–1.79) | 0.014 |
|  | Eastern | 4.9 | 0.35 (0.24–0.53) | 0.69 (0.47–1.02) | 0.065 |
|  | Luapula | 5.1 | 0.37 (0.22–0.63) | 0.65 (0.39–1.10) | 0.109 |
|  | North-Western | 4.7 | 0.34 (0.22–0.54) | 0.53 (0.35–0.82) | 0.004 |
|  | Northern | 9.8 | 0.75 (0.56–0.99) | 1.37 (1.01–1.85) | 0.045 |
|  | Southern | 4.6 | 0.33 (0.22–0.50) | 0.53 (0.36–0.76) | <0.001 |
|  | Western | 5.4 | 0.39 (0.26–0.58) | 0.66 (0.45–0.96) | 0.029 |
| Education | No education | 8.7 | Ref | Ref | - |
|  | Primary | 6.9 | 0.77 (0.57–1.04) | 0.77 (0.57–1.04) | 0.091 |
|  | Secondary | 9.0 | 1.04 (0.76–1.43) | 0.78 (0.56–1.07) | 0.117 |
|  | Higher | 17.2 | 2.18 (1.50–3.15) | 0.78 (0.53–1.15) | 0.212 |
| Wealth quintile | Poorest | 4.9 | Ref | Ref | - |
|  | Poorer | 4.9 | 1.00 (0.78–1.28) | 0.92 (0.72–1.19) | 0.538 |
|  | Middle | 6.5 | 1.35 (1.03–1.77) | 0.98 (0.72–1.34) | 0.912 |
|  | Richer | 10.3 | 2.23 (1.70–2.91) | 1.24 (0.88–1.77) | 0.222 |
|  | Richest | 14.7 | 3.32 (2.56–4.32) | 1.60 (1.08–2.39) | 0.020 |
| Marital status | Never married | 7.1 | Ref | Ref | - |
|  | In union | 8.5 | 1.22 (1.00–1.49) | 0.65 (0.52–0.81) | <0.001 |
|  | Formerly in union | 14.1 | 2.14 (1.70–2.69) | 1.05 (0.81–1.37) | 0.713 |
| Employment | Not employed | 6.1 | Ref | Ref | - |
|  | Employed | 11.9 | 2.08 (1.80–2.41) | 1.43 (1.22–1.68) | <0.001 |
| Any weekly media exposure | No | 6.8 | Ref | Ref | - |
|  | Yes | 10.4 | 1.61 (1.39–1.86) | 0.97 (0.82–1.15) | 0.760 |

### What a conventional analysis would report

Read at face value, the three rounds carrying a direct item show a substantial decline: among men, current drinking falls from 42.0% (95% CI 39.9–44.1) in 2007 to 37.6% (95% CI 36.3–39.0) in 2013–14 and 28.2% (95% CI 27.0–29.3) in 2024; among women, 11.1% (95% CI 9.9–12.4), 10.4% (95% CI 9.5–11.3) and 8.8% (95% CI 8.0–9.6). A conventional analysis would publish these; here they are the claim under test. Behind the men’s series lie two changes of instrument and one of beverage frame, the women’s carrying a changed reference period.

### Nesting bounds

Because the 2013–14 round carries both an undated status item and a count of drinking days in the last week, the thirty-day prevalence it would have recorded must lie between the two. For men that bracket is 30.5% to 37.6% against an observed 2024 value of 28.2%; for women, 6.1% to 10.4% against 8.8%. Here the men’s change is signed, the 2024 value falling below the whole bracket and implying a decline of 2.3 to 9.5 percentage points. The female value sits inside its bracket, so no sign attaches: decline, no change, or a modest rise all remain consistent with the data.

### Season matching, and where the men’s decline dissolves

Restricting both rounds to their shared fieldwork months removes that result. The 2013–14 round was in the field from August to December and January to April, the 2024 round from January to July, leaving January to April as their only common stretch. Season matters because prevalence defined over a recent window moves with the calendar as an undated status item cannot, so comparing unmatched parts of the year confounds season with trend.

On the common months alone, the male bracket moves to 26.9% to 35.8% against an observed 28.1%. The 2024 value now falls within the bracket and the change is no longer signed, lying between -7.6 and +1.2 percentage points; for women it remains unsigned, between -0.2 and +3.7. The unadjusted male comparison had cleared its bracket by only 2.3 percentage points, less than the 3.6-point seasonal movement in the 2013–14 seven-day figure itself.

### The lifetime analogue

A third, independent test draws on the 2024 item’s separation of never drinkers from those who did not drink in the past month. Lifetime consumption bounds current status from above, allowing comparison with the 2013–14 status item. Across all fieldwork months this implies a male decline of at least 2.5 percentage points; on common months it shrinks to 0.6, well inside sampling error. For women no decline is identified at all. Two independent strategies agree with the first: the change cannot be signed.

### An instrument-constant series

The partner items give the one series whose wording holds constant across rounds. Among ever-married women, the proportion reporting that their partner drinks alcohol fell from 53.7% (51.7–55.6) in 2007 to 46.5% (44.9–48.1) in 2013–14 and 37.7% (35.5–39.9) in 2018, then barely moved, at 37.0% (35.4–38.6) in 2024. Partner drunkenness followed the same path: 19.9% (18.3–21.5), 15.3% (14.2–16.4), 12.7% (11.6–13.8) and 12.9% (11.8–13.9).

As segment slopes, partner drinking fell by 1.19 percentage points a year between 2007 and 2013–14 and by 1.75 between 2013–14 and 2018, but by only 0.12 thereafter; for partner drunkenness the final segment is +0.03 percentage points a year, indistinguishable from flat. Shape matters more than level here. A straight line is formally rejected for both (Q = 15.9 and 14.7 on 2 degrees of freedom; p = 0.0003 and 0.0006), so constant annual change, the assumption behind any linear extrapolation, does not describe these data.

Composition can be ruled out: the pattern holds after direct standardisation to the 2024 age distribution, after restriction to women currently in union, and separately within urban and rural areas. Module non-completion among selected women ranged from 0.4% to 2.0% across rounds and item non-response never exceeded 0.3%, neither large enough to produce a movement of this size.

### Does the proxy measure what it claims?

Constant wording does not by itself guarantee that a series measures the same thing over time, so the measurement properties were tested directly. Linking each module-eligible woman to her co-resident husband gives 2,683 couples in 2007, 6,092 in 2013–14 and 4,555 in 2024. Among husbands who reported drinking, the proportion whose wives also reported them as drinkers fell from 86.0% to 79.3% and then to 66.8%; specificity held between 83.9% and 88.2%.

Sensitivity alone fell. Wives increasingly fail to report drinking their husbands themselves report, while remaining just as accurate about husbands who do not drink, so part of the proxy’s decline is instrument decay, not behaviour. Drifting in the same direction as the direct measure, it cannot show that decline to be genuine. It still counts against the hypothesis that the 2024 change in question wording created the drop; a general shift in willingness to report drinking, moving both series together, lies beyond what it can exclude.

The decay works in the direction of decline. If the instrument is pushing the series downwards, a measured flattening after 2018 is a conservative reading, and the plateau survives it. Much remains unsettled, though nothing here supports the view that alcohol use continued falling over the most recent period.

### What is identified, and what is not The 2024 cross-section

The comparability problem does not arise for the 2024 estimates: current drinking is reported by 28.2% (95% CI 27.0–29.3) of men and 8.8% (95% CI 8.0–9.6) of women. Of men, 64.9% have never consumed alcohol and 6.9% have consumed but not in the past month; among women, 84.5% and 6.7%. That structure constrains any projection: continued decline in current drinking would require lifetime abstention and cessation together to reach levels far above those observed.

Men who drink reported a mean of 7.2 drinking days in the past month and 5.5 drinks on a usual drinking day, with 41.3% reporting a usual heavy quantity of five or more drinks. Women who drink reported 5.1 days and 3.7 drinks, with 37.1% at four or more. The sex difference lies mainly in whether people drink at all; intensity among drinkers is closer between the sexes.

Prevalence rises steeply with age in both sexes, from 5.2% among men aged 15–19 to 40.0% at 35–44, and from 2.8% to 13.3% among women.

The 2024-only multivariable models show a sex divergence that pooling conceals. Among women the wealth gradient is positive: relative to the poorest quintile, the adjusted odds of current drinking in the richest quintile are 1.60 (95% CI 1.08–2.39); among men it runs the other way, at 0.69 (95% CI 0.52–0.91). Urban residence raises the odds in both sexes, at 1.62 (95% CI 1.25–2.09) among women and 1.47 (95% CI 1.26–1.72) among men, while education shows no consistent independent association in either. Measures aimed at the commercial retail market will therefore reach a different population of women than of men.

#### Key findings

The argument of this paper is supported by five key findings, each of which is addressed in detail in the Discussion section.

The four rounds are not a comparable series. Across the period the alcohol item changes in three separate ways: the 2007 men’s question asks about beer alone, the 2024 question replaces an undated status item with a thirty-day recall, and the 2018 round carries no direct item at all. Measurement is therefore already confounded with behaviour in any between-round difference, before a single estimate is compared.

Neither sex’s change between 2013–14 and 2024 can be signed. Once the change is bounded on a common thirty-day basis, with both rounds restricted to the fieldwork months they share, the male bound runs from -7.6 to +1.2 percentage points; for women it runs from -0.2 to +3.7. Season matching removes the male decline that the unadjusted comparison appears to show.

The one instrument-constant series falls steeply and then stops. Partner-reported drinking declines from 53.7% in 2007 to 37.7% in 2018, and then holds at 37.0% in 2024. Segment slopes of -1.19 and -1.75 percentage points a year give way to -0.12; a constant-decline model is rejected (Q = 15.9, 2 df, p = 0.0003).

The proxy itself loses accuracy over time. Measured against husbands’ own reports in co-resident couples, the sensitivity of a wife’s report falls from 86.0% to 66.8%, with specificity holding between 83.9% and 88.2%. Because a third-party report drifts in the same direction as the self-report it might have been used to check, it cannot establish that the direct decline is genuine.

The 2024 cross-section is sound, with a wealth gradient that runs in opposite directions by sex. Current drinking is reported by 28.2% of men and 8.8% of women. With age, residence, province, education, marital status, employment and media exposure held constant, the odds in the richest quintile relative to the poorest are 1.60 (95% CI 1.08–2.39) among women and 0.69 (95% CI 0.52–0.91) among men; both sexes show an urban excess.

## Discussion

Three of the four identification strategies fail to yield a conclusive direction of change since 2013–14, whereas the fourth reveals a trajectory that plateaus after 2018, accompanied by an indicator whose measurement reliability diminishes over time. Collectively, this evidence precludes any affirmation of a national decline in alcohol use in Zambia. Notably, the 2024 cross-sectional data are robust to these methodological limitations, highlighting a persistent and substantial burden of alcohol use—especially among men and urban populations, with considerable quantities consumed per drinking occasion.

Four mechanisms, not mutually exclusive, plausibly contribute. The 2007 men’s item asked about beer alone, in a setting where traditional and illicitly distilled beverages account for most consumption [18]; that frame understates alcohol use in the round and flatters any subsequent fall. Reference period compounds the problem. Replacing an undated status question with a bounded thirty-day recall mechanically excludes infrequent drinkers, and retrospective windows depress reported quantity relative to prospective collection in the same people (32,33). Season enters because fieldwork periods do not overlap and a recent-window measure moves with the calendar (34). Reporting behaviour is the fourth: willingness to report a partner’s drinking has demonstrably declined in these data, raising the question of whether the same holds for one’s own.

Self-reported alcohol is among the least reliable of survey measures. Population surveys capture only a minority of the alcohol known to be sold (35), and validation against phosphatidylethanol in southern Africa found self-reported recent heavy drinking to have a sensitivity of approximately 20%, with under-reporting some three and a half times more likely among women than men (36). Optimal biomarker thresholds for detecting self-reported unhealthy drinking differ several-fold between African and North American studies, which the authors attribute partly to differential reporting (37). Reported levels in any round should be read as reported drinking, not drinking. The very low female estimates, on which the unresolved female trend rests, are also the most exposed to reporting distortion.

Discontinuities of this kind are not peculiar to alcohol or Zambia. Abrupt level shifts in repeated cross-sectional series have been traced to survey design and administration, not population change (38); two national surveys using identical wording produced prevalence estimates differing by a factor of 1.7 through sample composition, questionnaire context and non-response (39); and within the DHS itself methodological choices moved a prevalence estimate by one to three percentage points across rounds while internal patterns remained stable (40). Stable internal patterns are therefore no evidence of comparability.

It remains plausible that some of the observed changes reflect authentic shifts in alcohol consumption patterns; nevertheless, the available data do not establish whether alcohol use in Zambia has remained stable or fluctuated. Crucially, these data are insufficient to disentangle genuine trends from artefacts of measurement, rendering any point estimate that attributes the entire observed change to behavioural factors methodologically indefensible.

### Key findings set against evidence

Pronounced impact of survey instrument and question framing on prevalence estimates aligns with our observation that changing instruments across ZDHS rounds fundamentally limits comparability (41). While other research has modelled trends across changing data collection methods or harmonized protocols to enable long-term analysis (42–44), such adjustments are not feasible here due to data limitations and the absence of a bridging round. Accordingly, the gap in the 2018 round prevents direct estimation or cross-walking between instruments, echoing our conclusion that trend interpretation is fundamentally constrained in this context (45).

The influence of seasonality on alcohol use estimates, as demonstrated in our findings, is consistent with global research indicating substantial month-to-month fluctuations (46–48). Adjusting for season in our analysis eliminates the apparent male decline, mirroring results elsewhere that underscore the importance of restricting comparisons to overlapping fieldwork periods (49,50). These parallels reinforce our methodological approach and validate the decision to compare only the months shared between rounds, rather than applying assumed corrections.

The trajectory observed in our data, a marked decline followed by a plateau, finds a parallel in studies that also reject the assumption of constant rates of change (51,52). However, our results diverge from broader regional and global trends, some of which show persistent increases or decreases depending on the indicator and data source(52,53). These discrepancies highlight the limitations of our dataset, particularly the reliance on self-reported drinking among partnered men and the exclusion of unrecorded and illicit alcohol, which complicates direct comparison and reconciliation with external series (54,55).

Our finding that reporting behaviour changed over time, particularly the declining willingness to disclose a partner’s drinking, aligns with research on the social and methodological factors shaping survey responses (56–58). While third-party reports can deliver reliable estimates under certain conditions (59,60), the observed drift in willingness to report highlights the vulnerability of proxy measures to evolving social norms, which in turn challenges the reliability of trend analysis across survey rounds.

Our findings regarding socio-economic and urban–rural gradients in alcohol use are echoed and, at times, contradicted by international studies, with apparent disparities often hinging on the specific indicator or population examined (61–63). The complexity of these relationships is further underscored by the varying patterns in the literature, which reinforce our interpretation that the direction and magnitude of inequalities depend on whether prevalence, heavy episodic drinking, or other participation measures are considered. Thus, our cross-sectional evidence, showing pro-rich participation among women and a flatter gradient among men, resonates with certain studies while differing from others, reflecting the broader heterogeneity in global patterns.

### Implications for surveillance and policy

Monitoring the WHO target for the harmful use of alcohol from national survey data is, at present, beyond Zambia’s reach. The target is defined over consumption and heavy episodic drinking(64,65). However, the available ZDHS indicators measure participation; no round carries an occasion-based heavy-drinking item aligned to the WHO definition (66). The partner-drunkenness item is the closest available analogue, but it records a third-party perception, with no quantity or threshold attached and no reference period. Regional assessments of progress toward the target therefore rest on modelled series instead of national survey trends (53,67), and the same limitation affects attempts to evaluate whether particular policy instruments are working (68,69). Planning figures obtained by extrapolating these rounds are already in circulation and should be carefully examined.

Other survey programmes have shown what is achievable. Holding the same instrument constant across waves makes change interpretable, and harmonizing several surveys at the point of analysis, then triangulating them against an external consumption denominator, has yielded defensible national burden series for South Africa and for Germany. Item-level data-quality diagnostics run before a series is used for trend purposes, with published ranges in place of single figures, are already standard practice for other DHS indicators.

Despite these methodological constraints, the 2024 round retains significant value. It establishes a credible baseline and should be considered the foundation for a renewed sequence of surveillance.

**Figure 1.**
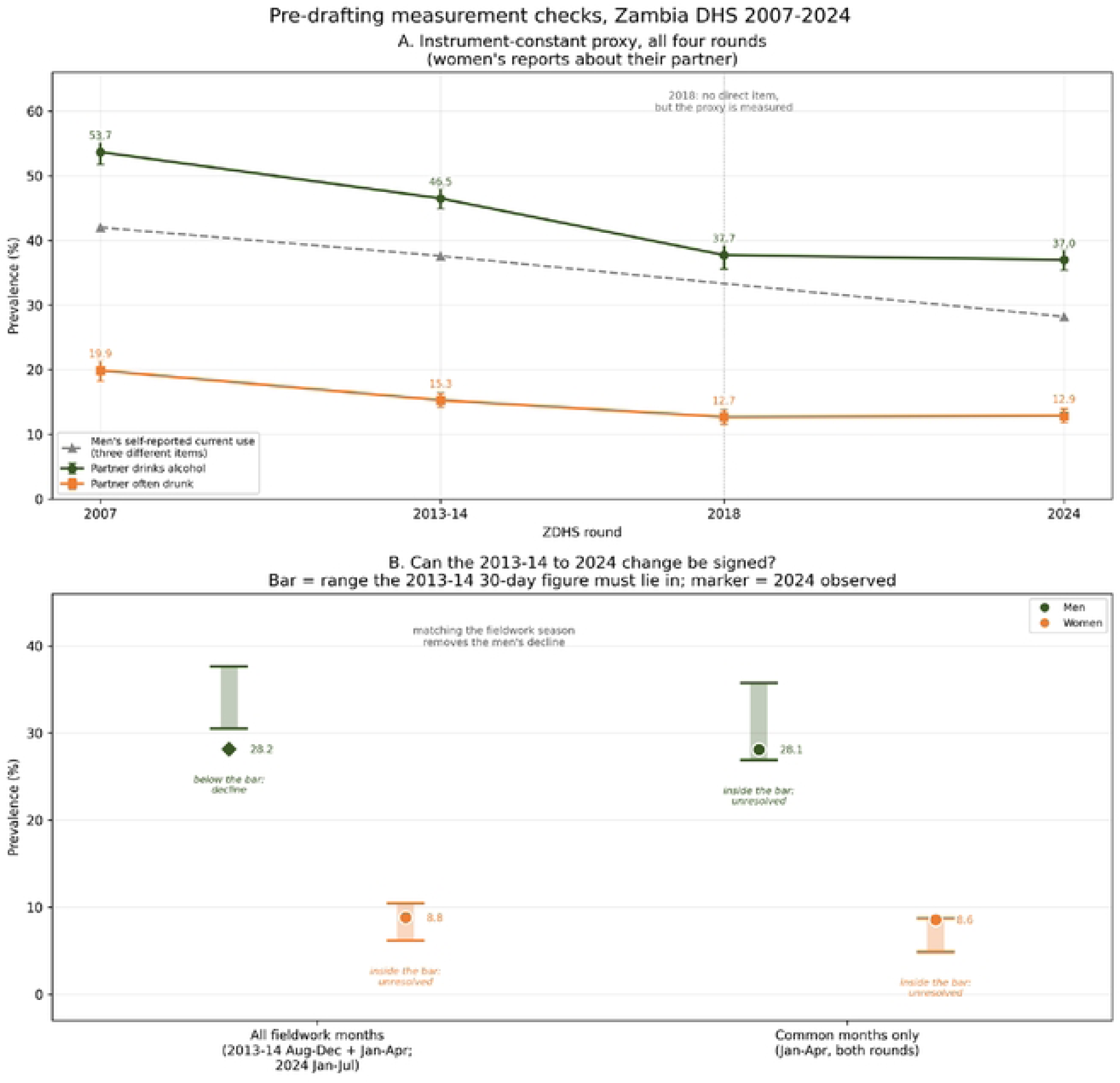
Top: the instrument-constant partner series across all four rounds, against the men’s self-reported series. Bottom: identification intervals for the 2013–14 to 2024 change, before and after restriction to common fieldwork months.

**Figure 2.**
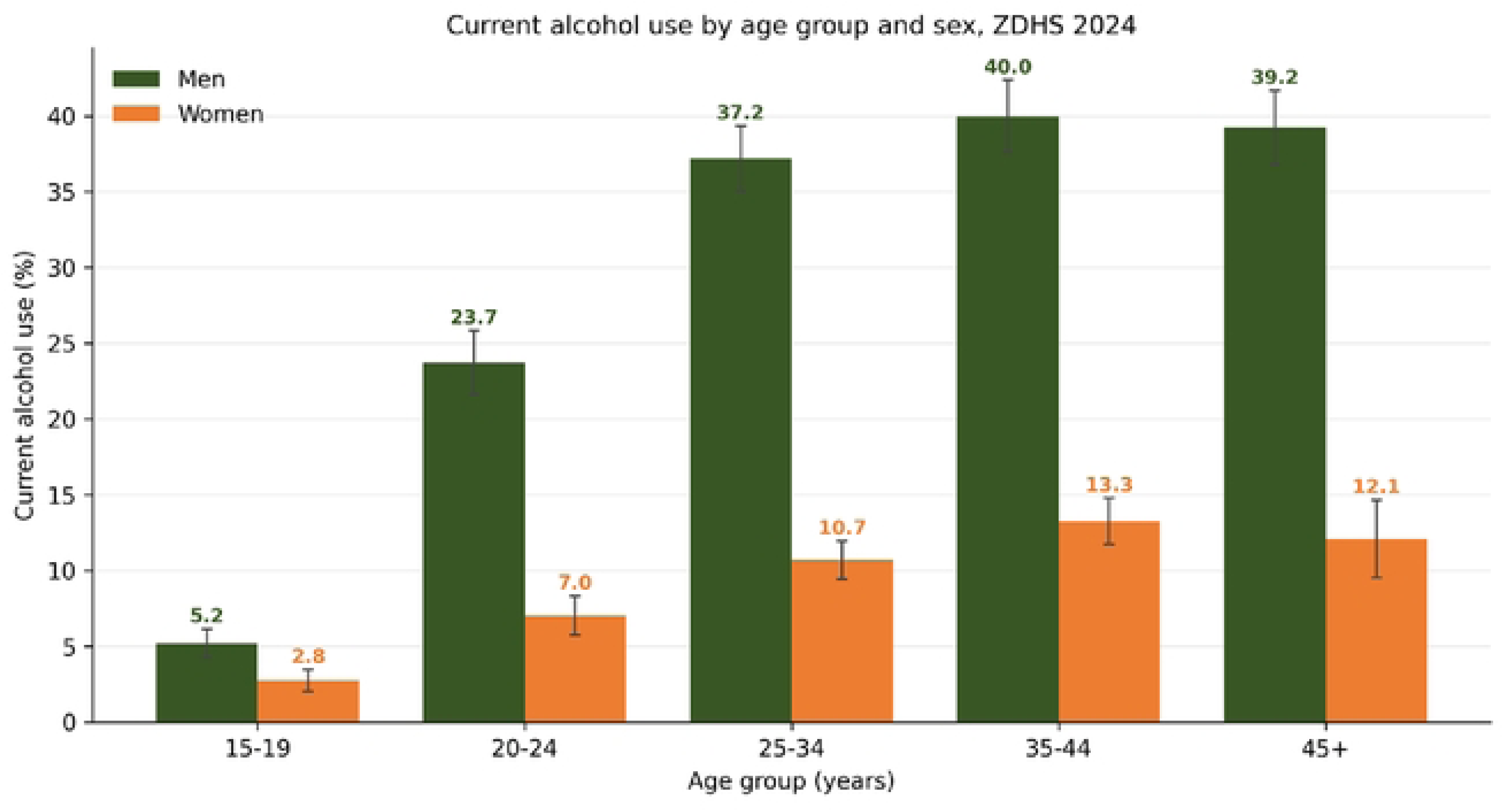
Current alcohol use by age band and sex, ZDHS 2024.

**Figure 3.**
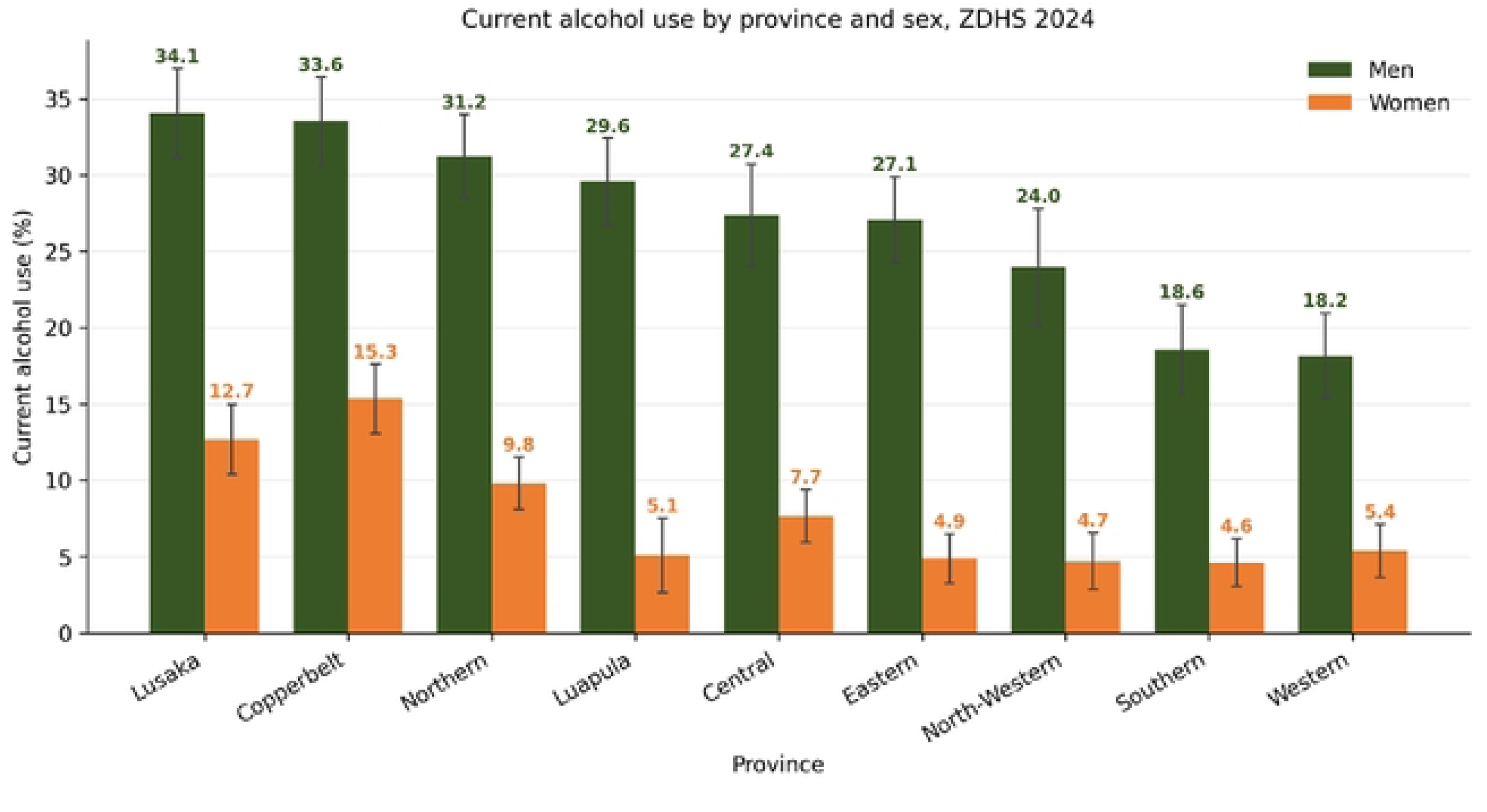
Current alcohol use by province and sex, ZDHS 2024.

## Conclusion

Retain the 2024 alcohol module unchanged in the next round. At this point consistency matters more than improvement to the instrument, since a further change would extend the period over which no trend can be measured. Add an occasion-based heavy-episodic-drinking item aligned to the WHO definition, which would make the indicator in which the target is written measurable. A validated brief screening instrument that already performs acceptably in Zambian populations would serve. Publish fieldwork month alongside estimates, and either balance fieldwork across seasons or publish season-adjusted figures. Retain the partner items. Where a couple’s sample exists, publication of the couple-linked concordance would replace an assumption about drift in the proxy with a measurement. Ask the alcohol module of both sexes on a universe that is held fixed across rounds. Triangulate survey estimates against an external consumption denominator. Recorded sales or excise data would allow a change in reporting to be distinguished from a change in drinking.

### Ethics approval

The study is a secondary analysis of publicly available, fully de-identified survey microdata; no individual can be identified from it and no participant was contacted. It was approved and authorized by the University of Zambia Biomedical Research Ethics Committee, approval number: **REF. NO. 8322-2026, Federal Assurance No. FWA00000338 IRB00001131 of IORG0000774 NHRAR-REC No 2021-05-0002**

## Data Availability

No legal or ethical concerns to address. The Study was approved. Approval number: REF. NO. 8322-2026, Federal Assurance No. FWA00000338 IRB00001131 of IORG0000774 NHRAR-REC No 2021-05-0002

## Author contributions

Shadreck Habbanti conceived the study idea, Picket Munkombwe conducted the Analysis, and the duo worked on the full paper. Cosmas Zyambo provided expert review

## Acknowledgements

The authors would like to acknowledgement the Zambia Central Statistics Office for conducting Demographic Health Surveys.

